# Epidemiological Study and Analysis of Factors Related to Skin Lesions Caused by Medical Disinfectants, and Personal Protective Equipment among Epidemic-Prevention Workers During the Severe Acute Respiratory Syndrome Coronavirus 2 Pandemic Lockdown Period

**DOI:** 10.1101/2024.08.15.24312034

**Authors:** Jing-Yi Hu, Xiu-Li Xiao, Yi Lu, Jian-Yong Su, Yan Zhang, Ting Shang, Chun-Hua Zhang, Lian Guo, Jian-Chao Wang

**Affiliations:** Chinese Medicine Department, Gaojing Town Community Health Service Center, 200439, Shanghai, China; Department of Dermatology, Shanghai Baoshan District Hospital of Integrated Traditional and Western Medicine, Shanghai University of Traditional Chinese Medicine, 200199, Shanghai, China; Department of Medical Cosmetology, Shanghai Baoshan District Hospital of Integrated Traditional and Western Medicine, Shanghai University of Traditional Chinese Medicine, 200199, Shanghai, China

**Keywords:** Prevalence, Risk factors, Skin lesions, Medical disinfectants, Personal protective equipment, SARS-CoV-2 Pandemic

## Abstract

**Objective:** The aim of this study was to investigate the prevalence of and potential risk factors associated with skin lesions resulting from the use of medical disinfectants and personal protective equipment (PPE) among epidemic prevention workers (including healthcare professionals, temporary sampling site workers, community members and volunteers) during the Severe Acute Respiratory Syndrome Coronavirus 2 (SARS-CoV-2) pandemic lockdown period in China.

**Methods:** We conducted a survey to investigate the prevalence and factors associated with skin lesions during SARS-CoV-2 pandemic lockdown period among epidemic prevention workers in Gaojing town of Baoshan distract, Shanghai, China.

**Results:** A total of 1033 questionnaires were reviewed, with 995 deemed valid. Among the 995 respondents, 209 (21.01%) reported comorbidities, while 786 (78.99%) were considered as controls. Autoimmune diseases, family history of dermatitis, cardiovascular diseases (CVDs), palmar and plantar hyperhidrosis, allergic diseases and the total time spent on skin cleansing and antisepsis procedures were identified as independent risk factors for these **s**kin lesions.

**Conclusion:** During the SARS-CoV-2 pandemic lockdown period, skin lesions among epidemic prevention workers was prevalent, which was primarily attributed to the use of medical disinfectants and PPE. These skin lesions frequently manifested as a combination of various subtypes across different areas of the body. Several individual factors, along with the total time spent on skin cleansing and skin antisepsis procedures, were identified as significant risk factors for the development of these skin lesions.

## 1. Introduction

During the SARS-CoV-2 pandemic, the occurrence of skin lesions resulting from medical disinfectants and personal protective equipment (PPE), such as device-related pressure injuries (DRPIs), moisture-associated skin damage (MASD) and skin tears (STs), significantly affected the physical and mental well-being of individuals involved in epidemic-prevention efforts. Early domestic studies published in 2020 revealed that the incidence of these skin lesions ranged from 28.44% to 93.39% ^[3-4]^. DRPIs incidence ranged from 26.58% to 57.51% ^[1-5]^. In the literature prior to 2019, the incidence of STs ranged from 1.06% to 22.00% ^[8-9]^, while the incidence of MASD was 8.83% ^[5]^. Additionally, the rates of dermal impregnation and allergic contact dermatitis have increased to 62.42% and 58.91% ^[2]^, respectively.

Previous studies have exclusively focused on medical personnel between the ages or twenty and forty years in SARS-CoV-2-designated hospitals in China ^[2-5]^. However, during the SARS-CoV-2 pandemic lockdown period (January 1st, 2022 to March 31st, 2023), a substantial number of SARS-CoV-2 nucleic acid detection samples were collected during the outbreak of SARS-CoV-2 infections. Healthcare professionals, temporary sampling site workers, community members and volunteers made significant effort to combat the SARS-CoV-2 outbreak. During the period spanning from December 2019 to January 2022, there was a noticeable shift in public health priorities, with a greater emphasis on quarantine measures and prevention strategies rather than solely treating and controlling the epidemic. The increased demand for community-based prevention support has led to the active involvement of various community resources (including nonhealth-care workers) in epidemic prevention efforts within urban communities in China. However, further investigation is required to determine whether the prevalence and characteristics of skin lesions has changed over time. Furthermore, the literature has focused primarily on factors such as occupation, workforce, age, sex and protection level ^[3-4]^, while personal factors such as autoimmune or allergic diseases in relation to disease onset remain unexplored. Therefore, the findings of this research will contribute to the clinical understanding of this issue.

## 2. Materials and methods

### 2.1. Research subjects

Between 1st April 2023 and 1st August 2023, a cross-sectional online survey was conducted in Gaojing town, Baoshan district, Shanghai. The recruitment of potential participants was facilitated through a network sampling approach, where the Ministry of Public Health and local government agencies were contacted to distribute a link to the online survey to all community neighborhood committees in Gaojing town and the health facilities situated in and around the aforementioned area. Healthcare professionals, temporary sampling site workers, community members and volunteers who were engaged Level 1-3 barrier protection measures to prevent the transmission of SARS-CoV-2 were recruited on a voluntary basis. No limitations based on age, sex or nationality were imposed. The exclusion criteria included incomplete or inaccurate questionnaire responses, as well as data that could not be effectively analyzed or appropriately characterized.

### 2.2. Study design, data collection, and sample size

This study adopted a retrospective cross-sectional design. A survey was conducted using a self-administered questionnaire and the reliability and validity of the presurvey data, which exceeded 0.9, were analyzed. Data collection was facilitated through the “Sojump” website and background system. Following data collection, incomplete or duplicate questionnaires were removed by information technicians, and a database was established.

The sample size calculation was based on the calculation for estimating prevalences in cross-sectional studies (*n*=Z_σ_^2^ × *pq/d*^2^). The incidence of the surveyed population was estimated with 95% confidence using Zσ, which was determined to be 1.96. Previous literature indicated an approximate incidence of 50% for the skin lesions evaluated, resulting in *p*=50% and *q*=1-*p*=50%. The allowable error, d, was estimated to be 0.1 times *p*, or 5%. Consequently, the sample size, *n*, was determined to be 387 for each group (the case group and the control group), resulting in a total sample size of 774. Accounting for an expected invalid response rate of 10%, the total sample size was adjusted to 847.

A total of 1033 questionnaires were recovered and collected, 31 incomplete questionnaires were excluded, and 7 duplicate questionnaires were obtained, resulting in 995 valid questionnaires, yielding a 96.3% completion rate.

### 2.3. Measurements

Statistical analyses were conducted using SPSS software (version 25.0). Normally distributed data are presented as 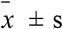. Comparisons between two groups were performed using independent sample t-tests. The frequency was used to describe the count data.

The χ^2^ test was employed to analyze the difference in prevalence between the two groups. A logistic regression analysis model was used to examine potential risk factors for the occurrence of these skin lesions, utilizing a significance level of alpha=0.05. Nonnormally distributed data are presented as the median (M), and 25th and 75th percentiles (P25 and P75, respectively). The Mann-Whitney U test was utilized to compare the two groups. Results for which P < 0.05 were determined to be statistically significant.

## 3. Results

### 3.1.1. Baseline data distribution of the survey population

The baseline data of the 995 subjects, including sex, age, occupation, workforce and protection level, are shown in Table 1. Of the participants, 23.51% were male. All participants were aged 18-80 years, and the mean age of all participants was 42.98±12.46 years.

**Table 1.**
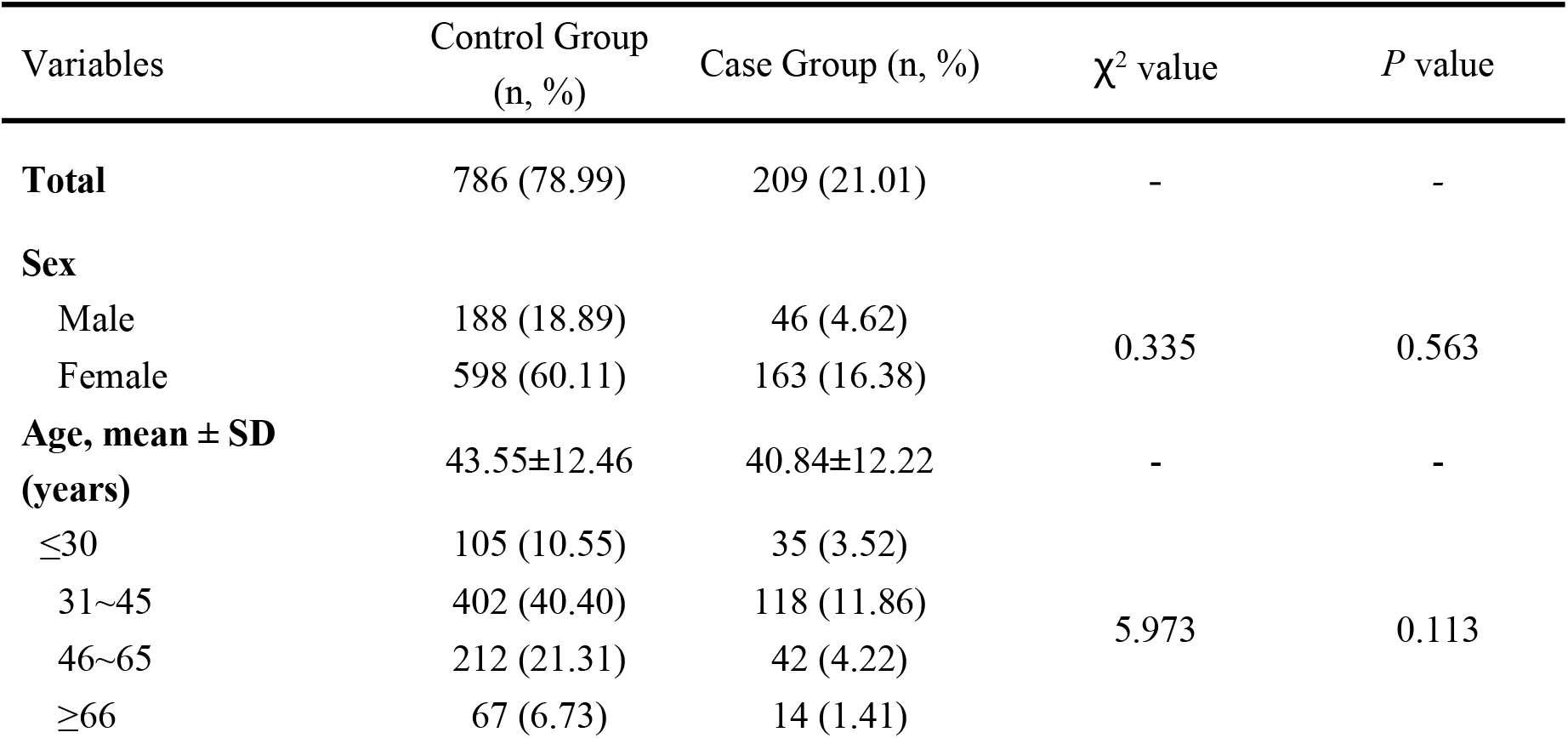
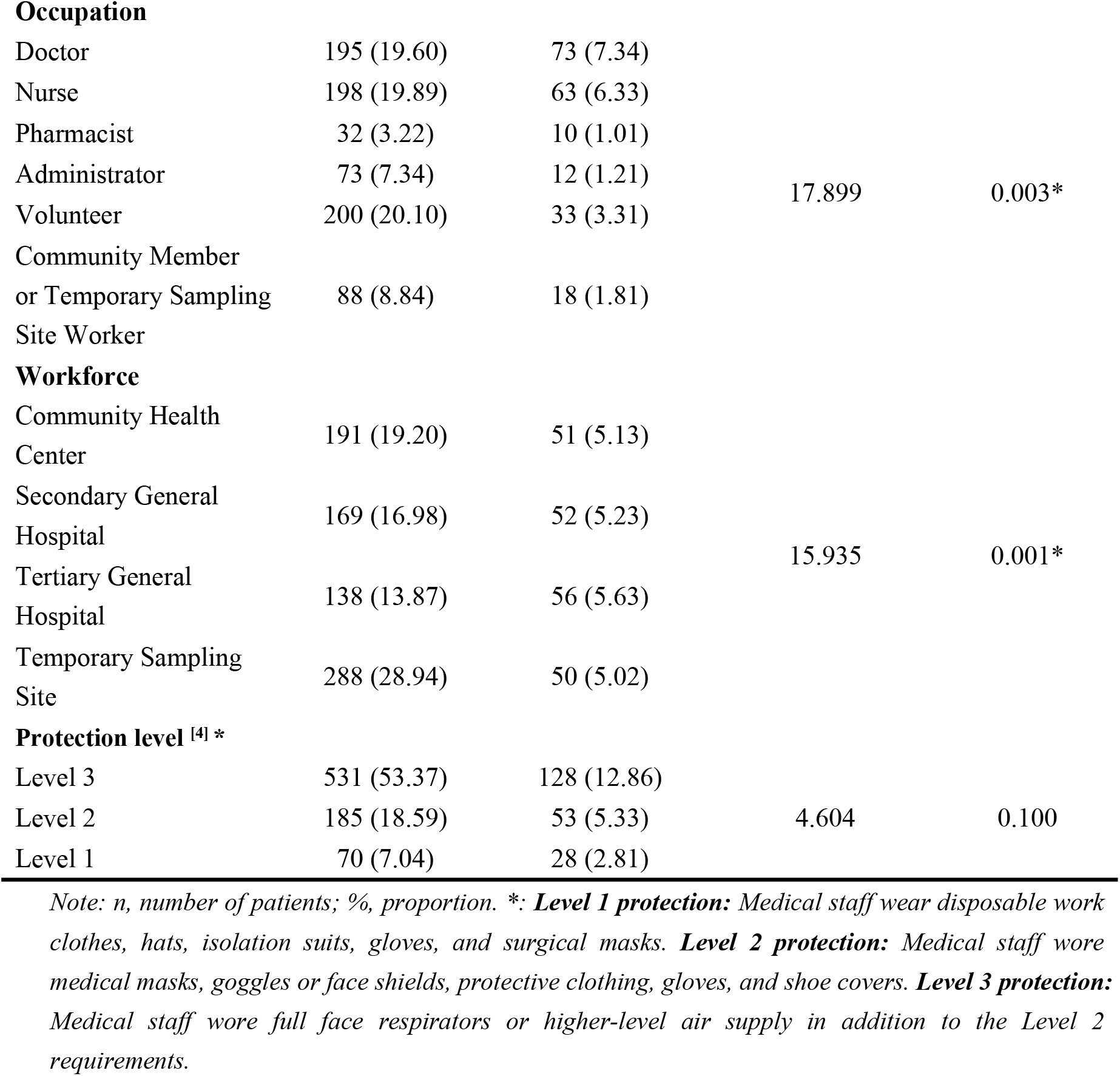
Baseline data distribution of the survey population (n=995)

| Variables | Control Group<br>(n, %) | Case Group (n, %) | $\chi^2$ value | $P$ value |
| --- | --- | --- | --- | --- |
| **Total** | 786 (78.99) | 209 (21.01) | - | - |
| **Sex** |  |  |  |  |
| Male | 188 (18.89) | 46 (4.62) | 0.335 | 0.563 |
| Female | 598 (60.11) | 163 (16.38) |  |  |
| **Age, mean ± SD**<br>**(years)** | 43.55±12.46 | 40.84±12.22 | - | - |
| $\leq 30$ | 105 (10.55) | 35 (3.52) | 5.973 | 0.113 |
| 31~45 | 402 (40.40) | 118 (11.86) |  |  |
| 46~65 | 212 (21.31) | 42 (4.22) |  |  |
| $\geq 66$ | 67 (6.73) | 14 (1.41) | | |
| **Occupation** |  |  |  |  |
| Doctor | 195 (19.60) | 73 (7.34) | 17.899 | 0.003* |
| Nurse | 198 (19.89) | 63 (6.33) |  |  |
| Pharmacist | 32 (3.22) | 10 (1.01) |  |  |
| Administrator | 73 (7.34) | 12 (1.21) |  |  |
| Volunteer | 200 (20.10) | 33 (3.31) |  |  |
| Community Member<br>or Temporary Sampling<br>Site Worker | 88 (8.84) | 18 (1.81) |  |  |
| **Workforce** |  |  |  |  |
| Community Health<br>Center | 191 (19.20) | 51 (5.13) | 15.935 | 0.001* |
| Secondary General<br>Hospital | 169 (16.98) | 52 (5.23) |  |  |
| Tertiary General<br>Hospital | 138 (13.87) | 56 (5.63) |  |  |
| Temporary Sampling<br>Site | 288 (28.94) | 50 (5.02) |  |  |
| **Protection level <sup>[4]</sup> \*** |  |  |  |  |
| Level 3 | 531 (53.37) | 128 (12.86) | 4.604 | 0.100 |
| Level 2 | 185 (18.59) | 53 (5.33) |  |  |
| Level 1 | 70 (7.04) | 28 (2.81) |  |  |

*Note: n, number of patients; %, proportion. \*: **Level 1 protection:** Medical staff wear disposable work* *clothes, hats, isolation suits, gloves, and surgical masks. **Level 2 protection:** Medical staff wore* *medical masks, goggles or face shields, protective clothing, gloves, and shoe covers. **Level 3 protection:*** *Medical staff wore full face respirators or higher-level air supply in addition to the Level 2* *requirements.*

The distributions of sex, age and proportions related to occupation, workforce and protection level were tested using the chi-square test. There were no significant differences in sex, ages, or protection level between the two groups, however, we noted significant differences in the proportions of occupations (*χ*^2^=17.899, *P*=0.003) and workers (*χ*^2^=15.935, *P*=0.001) between the two groups.

### 3.1.2. Personal factors and other potential risk factors

Differences in personal factors and other potential risk factors, such as variations in the amount of exposure to disinfectants, the total time spent on skin cleansing and skin antisepsis procedures, the total time spent on wearing PPE and the protection level, were tested using the chi-square test (Table 2).

**Table 2.**
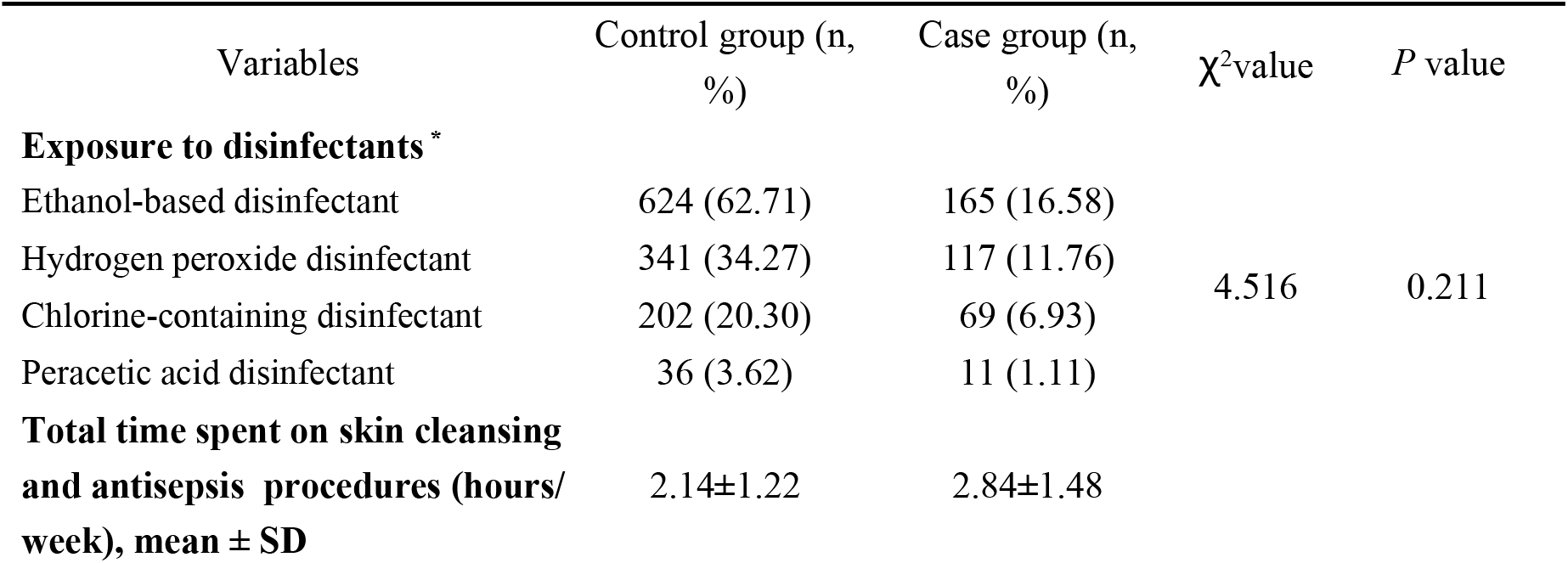
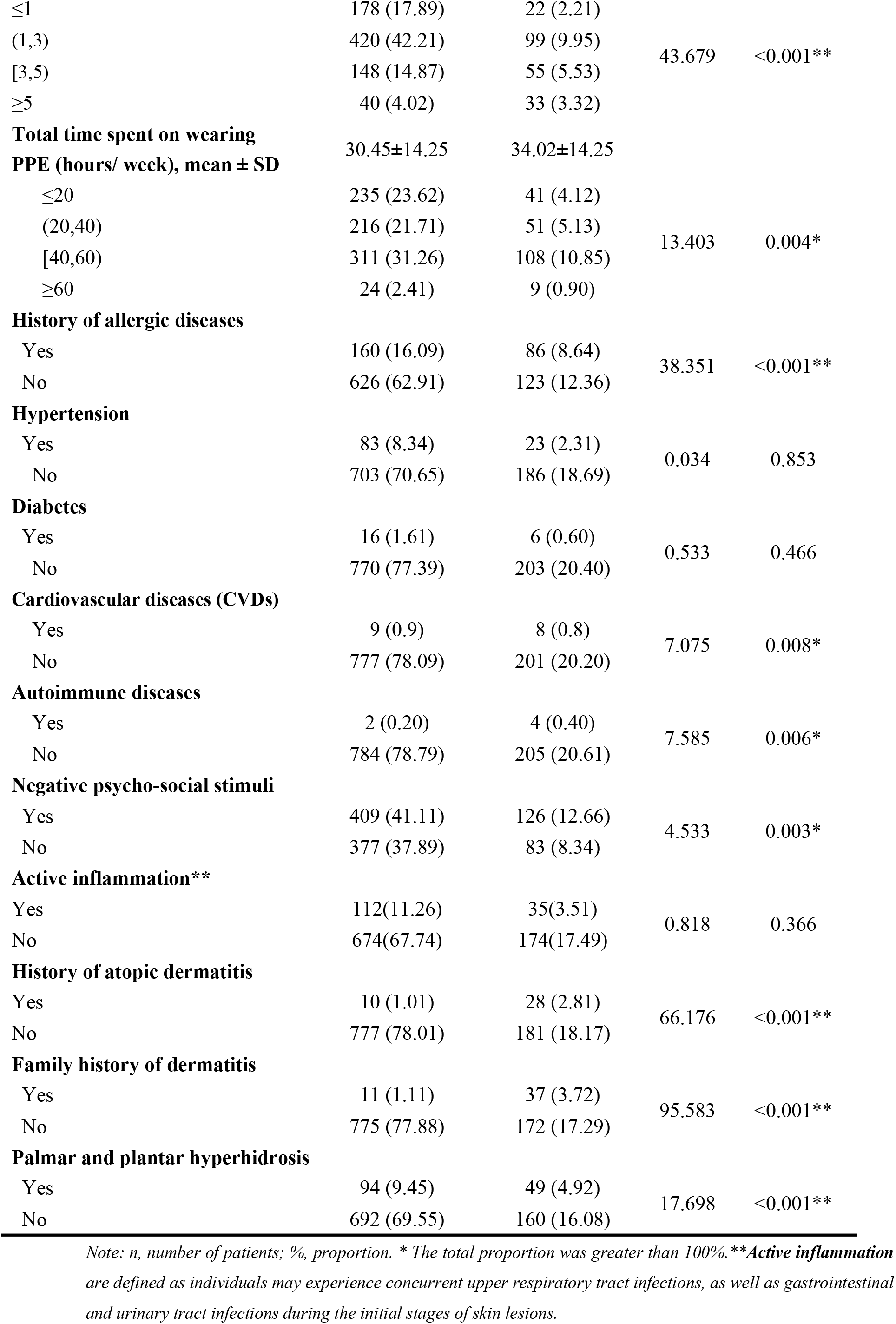
Distribution of related personal factors among participants (n=995)

| Variables | Control group (n,<br>%) | Case group (n,<br>%) | $\chi^2$value | *P* value |
| --- | --- | --- | --- | --- |
| **Exposure to disinfectants \*** |  |  |  |  |
| Ethanol-based disinfectant | 624 (62.71) | 165 (16.58) | 4.516 | 0.211 |
| Hydrogen peroxide disinfectant | 341 (34.27) | 117 (11.76) |  |  |
| Chlorine-containing disinfectant | 202 (20.30) | 69 (6.93) |  |  |
| Peracetic acid disinfectant | 36 (3.62) | 11 (1.11) |  |  |
| **Total time spent on skin cleansing**<br>**and antisepsis procedures (hours/**<br>**week), mean ± SD** | 2.14±1.22 | 2.84±1.48 | | |
| ≤1 | 178 (17.89) | 22 (2.21) | 43.679 | <0.001** |
| (1,3) | 420 (42.21) | 99 (9.95) |  |  |
| [3,5) | 148 (14.87) | 55 (5.53) |  |  |
| ≥5 | 40 (4.02) | 33 (3.32) |  |  |
| **Total time spent on wearing PPE (hours/ week), mean ± SD** | 30.45±14.25 | 34.02±14.25 |  |  |
| ≤20 | 235 (23.62) | 41 (4.12) | 13.403 | 0.004* |
| (20,40) | 216 (21.71) | 51 (5.13) |  |  |
| [40,60) | 311 (31.26) | 108 (10.85) |  |  |
| ≥60 | 24 (2.41) | 9 (0.90) |  |  |
| **History of allergic diseases** |  |  |  |  |
| Yes | 160 (16.09) | 86 (8.64) | 38.351 | <0.001** |
| No | 626 (62.91) | 123 (12.36) |  |  |
| **Hypertension** |  |  |  |  |
| Yes | 83 (8.34) | 23 (2.31) | 0.034 | 0.853 |
| No | 703 (70.65) | 186 (18.69) |  |  |
| **Diabetes** |  |  |  |  |
| Yes | 16 (1.61) | 6 (0.60) | 0.533 | 0.466 |
| No | 770 (77.39) | 203 (20.40) |  |  |
| **Cardiovascular diseases (CVDs)** |  |  |  |  |
| Yes | 9 (0.9) | 8 (0.8) | 7.075 | 0.008* |
| No | 777 (78.09) | 201 (20.20) |  |  |
| **Autoimmune diseases** |  |  |  |  |
| Yes | 2 (0.20) | 4 (0.40) | 7.585 | 0.006* |
| No | 784 (78.79) | 205 (20.61) |  |  |
| **Negative psycho-social stimuli** |  |  |  |  |
| Yes | 409 (41.11) | 126 (12.66) | 4.533 | 0.003* |
| No | 377 (37.89) | 83 (8.34) |  |  |
| **Active inflammation\*\*** |  |  |  |  |
| Yes | 112(11.26) | 35(3.51) | 0.818 | 0.366 |
| No | 674(67.74) | 174(17.49) |  |  |
| **History of atopic dermatitis** |  |  |  |  |
| Yes | 10 (1.01) | 28 (2.81) | 66.176 | <0.001** |
| No | 777 (78.01) | 181 (18.17) |  |  |
| **Family history of dermatitis** |  |  |  |  |
| Yes | 11 (1.11) | 37 (3.72) | 95.583 | <0.001** |
| No | 775 (77.88) | 172 (17.29) |  |  |
| **Palmar and plantar hyperhidrosis** |  |  |  |  |
| Yes | 94 (9.45) | 49 (4.92) | 17.698 | <0.001** |
| No | 692 (69.55) | 160 (16.08) |  |  |

*Note: n, number of patients; %, proportion. \* The total proportion was greater than 100%. \*\*Active inflammation* *are defined as individuals may experience concurrent upper respiratory tract infections, as well as gastrointestinal* *and urinary tract infections during the initial stages of skin lesions.*

Allergic diseases are systemic disorders caused by an impaired immune system. The total number of patients in the case group was 86, including those with allergic dermatitis (n=31), allergic rhinitis (n=35) and bronchial asthma (n=20).The total number of patients in the control group was 160, including those with allergic dermatitis (n=10), allergic rhinitis(n=138) and bronchial asthma (n=12).

Cardiovascular diseases(CVDs) are a collective term for heart and vascular diseases, mainly including coronary heart disease, cerebrovascular diseases (such as stroke), peripheral arterial diseases, congenital heart disease, deep vein thrombosis, and pulmonary embolism.The total number of patients in the case group was 8, including those with cerebral infarction (n=4) and coronary artery disease (n=4). The total number of patients in the control group was 9, including coronary artery disease (n=9).

Autoimmune diseases are defined as an abnormal response of B cells and/or T cells towards endogenous antigens, which in turn leads to self-directed immunity and eventually presents as either localized tissue damage or as a systemic disease.The total number of patients in the case group was 4, which included patients with Sjögrens syndrome (n=1), Hashimoto’s thyroiditis (n=1), IgA nephropathy (n=1) and psoriatic arthritis (n=1). There were 2 patients in the control group is, one with IgA nephropathy (n=1) and one with Hashimoto’s thyroiditis(n=1).

Negative Psychosocial Stimuli: The total number of patients in the case group was 126, including those with stress (n=26), insomnia (n=33),depression (n=29) and anxiety (n=38).There were 409 individuals in the control group, including those with stress(n=89), insomnia (n=73),depression (n=98) and anxiety (n=149). Family history of dermatitis: The total number of patients in the case group was 37, including those with atopic dermatitis (n=31), psoriasis (n=4), exfoliative keratosis (n=1) and seborrheic dermatitis (n=1). There were 11 individuals in the control group, including those with atopic dermatitis (n=10) and seborrheic dermatitis (n=1).

There were no significant differences in exposure to disinfectants and the prevalence of hypertension, diabetes or active inflammation between the two groups. However, we noted significant differences between the two groups in the total time spent on skin cleansing and skin antisepsis procedures (*χ*^2^=43.679, *P*<0.001) and the total time spent on wearing PPE (*χ*^2^=13.403, *P*=0.004). Furthermore, the prevalence of these skin lesions among participants with allergic diseases was significantly greater than that among participants without allergic diseases (*χ*^2^=38.351, *P*<0.001). The prevalence of these skin lesions among participants with cardiovascular diseases, autoimmune diseases, a history of atopic dermatitis, a family history of dermatitis and palmar & plantar hyperhidrosis was also greater than that among participants without diseases (All P<0.05). In addition, those who experienced negative psychosocial stimuli or a family history of dermatitis were more likely to have greater incidence of these skin lesions. (*χ*^2^=4.533, *P*=0.003 and *χ*^2^=95.583, *P*<0.001, respectively).

Of the 209 participants with comorbidities, 13 (31.0%) exhibited DRPIs, 165 (78.95%) had MASD, and twenty (9.57%) experienced both DRPIs and MASD. Additionally, one patient (0.48%) experienced both DRPIs and STs and another patient (0.48%) had both MASD and STs. Furthermore, nine participants (4.30%) presented with all three types of skin lesions.

### 3.2 Type-specific distribution patterns

There were 363 skin lesions in total, among which forty-three were DRPIs, and the head-face and neck areas were two of the most prevalent locations. Three hundred and seven out of 363 patients had MASD, the hand, neck and forearm and wrist area were the most vulnerable areas, in order of highest prevalence. Thirteen of 363 patients had STs, and the head-face area was the most common location (Table 3).

**Table 3.** Type-specific distribution patterns of skin lesions caused by medical disinfectants and personal protective equipment (n=363)

| Type of skin lesions | Count (n, %) | Scalp (n, %) | Head& face (n, %) | Neck (n, %) | Axillae (n, %) | Groin (n, %) | Forearm & wrist (n, %) | Dorsum of Hand (n, %) | Palm (n, %) | Ankle (n, %) | Foot (n, %) |
| --- | --- | --- | --- | --- | --- | --- | --- | --- | --- | --- | --- |
| DRPIs | 43(100) | 0(0) | 36 (83.72) | 7(16.28) | 0(0) | 0(0) | 0(0) | 0(0) | 0(0) | 0(0) | 0(0) |
| MASD | 307(100) | 14(4.56) | 17(5.54) | 26 (8.47) | 3(0.98) | 5(1.63) | 25(8.14) | 136 (44.30) | 69(22.48) | 7(2.27) | 5(1.63) |
| STs | 13(100) | 0(0) | 10 (76.93) | 1(7.69) | 0(0) | 1(7.69) | 0(0) | 0(0) | 0(0) | 1(7.69) | 0(0) |
| Total | 363(100) | 14(3.86) | 63 (17.36) | 34(9.37) | 3(0.83) | 6(1.65) | 25(6.88) | 136 (37.47) | 69(19.00) | 8(2.20) | 5(1.38) |

### 3.3 Distribution of onset time

This temporal analysis reveals two peaks between January 2022 and March 2023. The first peak occurred in June 2022, followed by a second larger peak from December 2022 through January 2023, which demonstrated that the early summer of 2022 and winter and early spring of 2023 were high-incidence seasons. This result is consistent with the fact that the lockdown was initiated in Shanghai in March 2022 and gradual reopening started in December 2022 (Figure 1).

**Figure 1.**
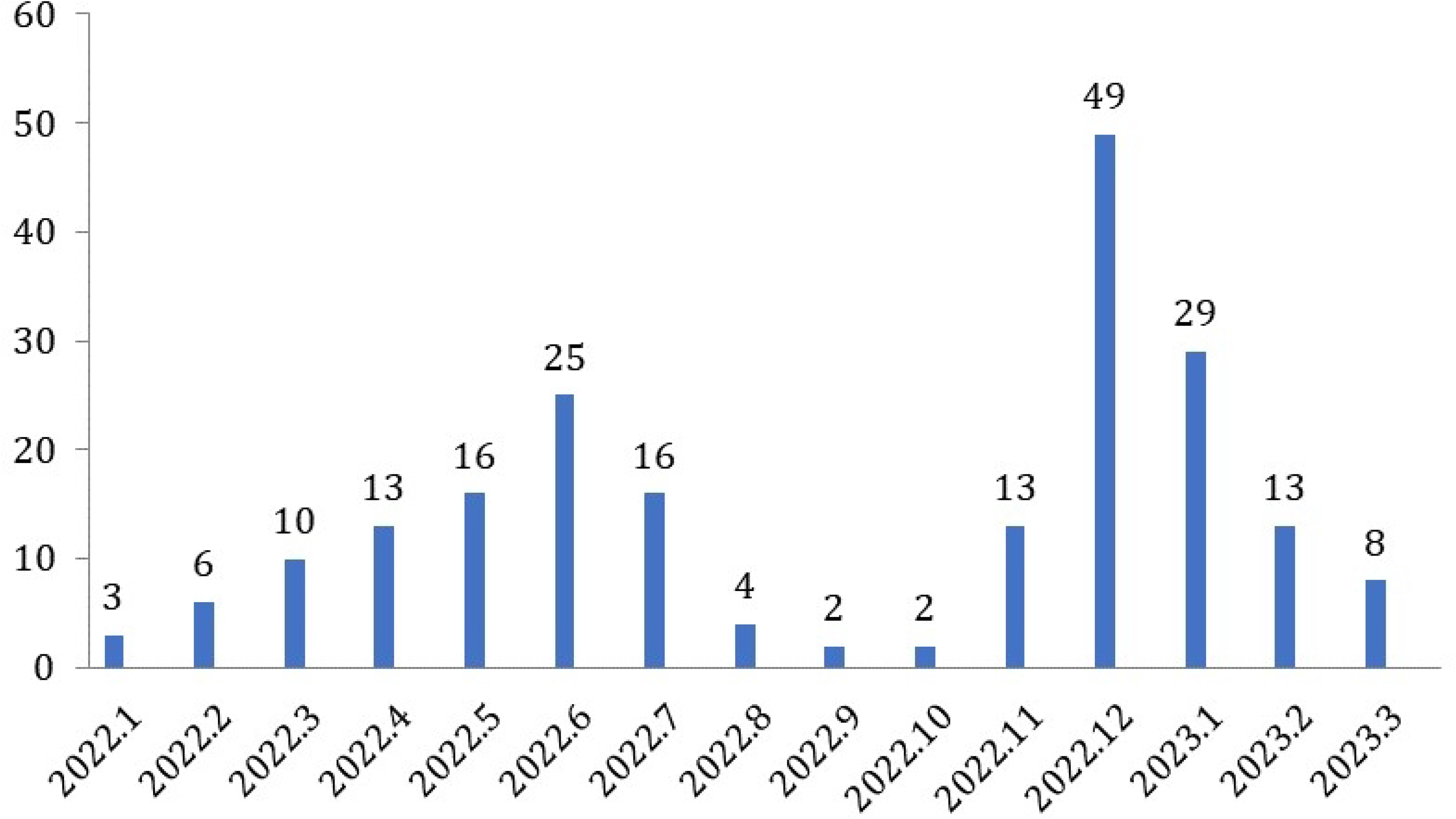
Temporal Distribution of the Onset of Skin Lesions Caused by Medical Disinfectants and Personal Protective Equipment (Histogram) (n=209)

### 3.2. Related factors

The potential independent variables included in logistic regression analysis are shown in Table 4. The variables “occupation”, “workforce”, “total time spent on wearing PPE”, “history of atopic dermatitis” and “negative psychosocial stimuli” were examined in the multivariate model, which revealed no statistically significant associations with the onset of skin lesions. Thus, the risk factors for the skin lesions examined included the total time spent on skin cleansing and antisepsis procedures, cardiovascular diseases, autoimmune diseases, allergic diseases, family history of skin dermatitis and palmar and plantar hyperhidrosis.

**Table 4.** Factors associated with skin lesions caused by medical disinfectants and personal protective equipment according to multivariate logistic regression.

| Variables | Multivariate |  |  |  |  |
| --- | --- | --- | --- | --- | --- |
|  | *B* | SE | OR | 95% CI | *P* value |
| **Occupation** |  |  |  |  | 0.656 |
| **Workforce** |  |  |  |  | 0.366 |
| **Total time spent on skin cleansing and antiseptic procedures (hours/ week)** |  |  |  |  | <0.0001** |
| ≤1 | -1.861 | 0.367 | 0.156 | (0.076,0.320) | <0.0001** |
| (1,3) | -1.099 | 0.292 | 0.333 | (0.188,0.591) | <0.0001** |
| [3,5) | -0.743 | 0.315 | 0.476 | (0.256,0.883) | 0.019 |
| ≥5 | 0.000 |  |  |  |  |
| **Total time spent on wearing PPE (hours/ week)** |  |  |  |  | 0.471 |
| **History of atopic dermatitis** | 1.302 | 1.192 | 3.677 | (0.355, 38.051) | 0.275 |
| **CVDs** | 1.643 | 0.596 | 5.173 | (1.610,16.624) | 0.006* |
| **Autoimmune diseases** | 2.434 | 1.076 | 11.405 | (1.383,94.034) | 0.024* |
| **Allergic diseases** | 0.563 | 0.192 | 1.755 | (1.205,2.556) | 0.003* |
| **Family history of skin dermatitis** | 3.563 | 1.130 | 35.255 | (3.850,322.845) | 0.002* |
| **Negative psychosocial stimuli** | 0.185 | 0.183 | 1.204 | (1.169, 2.823) | 0.311 |
| **Palmar and plantar hyperhidrosis** | 0.597 | 0.225 | 1.816 | (1.169,2.823) | 0.008* |

Note: \* $P < 0.05$ , \*\* $P < 0.001$ . SE, standard error. OR, odds ratio. 95% CI, 95% confidence interval.

The odds ratio(ORs) calculated were as follows: Cardiovascular diseases (OR=5.173; 95% CI=1.610 to 16.624; P=0.006), autoimmune diseases (OR=11.405; 95% CI=1.383 to 94.034; P=0.024), allergic diseases (OR=1.755; 95% CI=1.205 to 2.556; P=0.003), family history of skin dermatitis (OR=35.255; 95% CI=3.850 to 322.845; P=0.002) and palmar and plantar hyperhidrosis (OR=1.816; 95% CI=1.169 to 2.823; P=0.008). Factors associated with greater likelihood of suffering from skin lesions were the total time spent on skin cleansing and antisepsis procedures (P<0.001). Multivariable Logistic Regression demonstrated a statistically significant differences in total duration as follows: ≥5 hours and ≤1 hour (OR=0.156; 95% CI=0.076 to 0.320; P<0.001), 1-3 hours (OR=0.333; 95% CI=0.188 to 0.591; P<0.001), and 3-5 hours (OR=0.476; 95% CI=0.256 to 0.883; P=0.019), suggesting an increased risk of disease incidence with exposure.

## 4. Discussion

### 4.1 Characteristics and prevalence

Overall, surveys from 995 patients were examined and a chi-square test was used to compare demographic data between the two groups. The results revealed statistically significant differences in occupation and workforce, but no significant differences were observed in sex, age or protection level (P>0.05). These findings are in contrast with existing literature published between 2019 and 2020 ^[2-5]^, which primarily examined healthcare personnel between the ages of 20 to 40 years working in SARS-CoV-2-designated hospitals in China. These studies revealed unexpectedly high females to males ratios, along with a significant proportion of individuals using level 3 barrier PPE for protection and an imbalanced age distribution ^[2-5]^. However, during SARS-CoV-2 pandemic lockdown period, various stakeholders, including healthcare professionals, temporary sampling site workers, community members and volunteers, made substantial efforts to combat the SARS-CoV-2 virus. The disaggregation of data on sex and age became more detailed, and the distribution of protection levels became more equitable.

The prevalence of DRPIs and the proportion of participants exhibiting two or three types of skin lesions observed in the current study was less when compared to previous domestic literature covering the period from 2019 to 2020 ^[2-5]^, but align with earlier studies ^[1,6-8]^. Several potential hypotheses can be proposed to explain these results. First, it is plausible that the total duration of skin cleansing and PPE use among temporary sampling site workers, community members and volunteers was shorter than that among medical staff. This practice effectively mitigated the risk of developing DRPIs or the cooccurrence of other types of skin lesions, preventing simple skin lesions from becoming more severe. Second, the findings of this study exhibited a greater degree of generalizability when applied to a larger population. Third, variations in temperature, humidity and other climatic factors could account for the observed differences between these variables.

A total of 363 skin lesions were observed, forty-three of which were identified as DRPIs, primarily affecting the head-face and neck regions. The majority of the lesions (307 out of 363) were classified as MASD, with a higher prevalence observed in the hand and neck areas. Additionally, thirteen lesions were categorized as STs, predominantly occurring in the head-face region. These findings align with previous research studies ^[1-7]^. Notably, the occurrence of DRPIs lesions were primarily attributed to friction and sheer stress resulting from the use of nasal and facial masks or protective eyewear. Failure to address these stress-related issues may cause skin tearing. Hence, while STs are commonly associated with DRPIs, the two do not occur concurrently.

MASD occurs in regions characterized by excessive sweating and elevated levels of skin humidity, such as palms, soles, and axillae ^[6-7]^. Notably, this study revealed a greater incidence in the palms and head and face area compared to previous studies ^[6]^. This observation aligns with observations made in clinical practice. Since protective measures primarily include nasal and facial protection, as well as hand cleansing and disinfection, the vulnerability of the palms and head and face area is accentuated.

### 4.2 Analysis of related factors

Risk factors and skin lesions caused by medical disinfectants and PPE were identified through multivariable logistic regression. Autoimmune disease, a family history of skin dermatitis, CVDs, palmar and plantar hyperhidrosis, and allergic diseases were found to be independent risk factors for these **s**kin lesions. Variables mentioned above may be linked to genetic diversity and the immune microenvironment in vivo. However, these hypotheses need further confirmation. Therefore, in future research, we intend to increase the sample size and evaluate the levels of immune-inflammatory cells and immune-inflammatory factors, including tumor necrosis factor (TNF) and interleukin (IL).

The statistical analysis revealed that workforce, protection level, total time spent on wearing PPE, or psychosocial factors were not significantly associated with the onset of skin lesions (P>0.05).

Factors such as total duration of wearing PPE, history of atopic dermatitis and negative psychosocial stimuli might offer some protection, but further analysis revealed no significant difference between groups. Nevertheless, this speculation requires confirmation by additional studies with larger sample sizes.

However, the total time spent on skin cleansing and antisepsis procedures were identified as risk factors for these skin lesions. The findings of this study demonstrated a positive association between the duration of exposure and the ORs calculated, suggesting an increased risk of disease incidence. These estimates align with the observed patterns in real-world scenarios.

Occupation was not significantly associated with the onset of skin lesions. This finding differed from previous literature from 2019-2020 that studied healthcare workers in China^[2-5]^. It is speculated that the limited working hours of volunteers, community members or temporary sampling site workers might reduce the duration of exposure to medical disinfectants and PPE compared to that of medical staff, while doctors, nurses, pharmacists and administrators, might demonstrate a greater level of awareness regarding skin protection and are more likely to employ effective measures to prevent skin lesions. Therefore, occupation emerged as a confounding variable in relation to other factors within the framework of national epidemic prevention and control.

## 5. Conclusions

During the SARS-CoV-2 pandemic lockdown period, skin lesions among epidemic prevention workers was prevalent, which was primarily attributed to the use of medical disinfectants and PPE. These skin lesions frequently manifested as a combination of various subtypes across different areas of the body. The prevalence and characterization of these skin lesions varied over time.

Throughout the third year of the pandemic, SARS-CoV-2 and other pathogens circulated at relatively low levels. These skin lesions have the potential to persist long term and significantly impact quality of life. Drawing upon evidence from the literature, it is recommended that individuals who engage in skin cleansing and skin antisepsis protocols for extended periods, particularly after working in level three barrier protection for more than five hours, should take mandatory breaks or shifts.

Several individual factors, along with the total time spent on skin cleansing and antisepsis procedures, were identified as significant risk factors for the development of skin lesions. We suggest the use of a comprehensive screening scale to identify individuals at high risk for skin lesions, with the aim of implementing targeted interventions to prevent their occurrence. These interventions involve the early application of dressings, topical agents, and antiallergic medications to mitigate skin friction and moisture. Additionally, it is imperative to mobilize collective action in the field of public health to promote education on skin barrier protection as an essential preventive measure.

## Data Availability

All relevant data are within the manuscript and its Supporting Information files.

## Acknowledgment

Informed consent was obtained from all subjects involved in the study. This study was reviewed and approved by the Research Ethics Committee of Wusong Hospital, Zhongshan Hospital, Fudan University (IRB No.2022-SYY-15).

## Author contributions

Jing-Yi Hu, Xiu-Li Xiao: conceptualization, methodology, validation, investigation, data curation, writing-review and editing of the manuscript draft. Jing-Yi Hu: formal analysis, writing-original draft. Xiu-Li Xiao, Yi Lu, Jian-Yong Su: project administration. Jing-Yi Hu, Xiu-Li Xiao: funding acquisition. Xiu-Li Xiao: supervision. Yan Zhang, Ting Shang, Chun-Hua Zhang, Lian Guo and Jian-chao Wang: writing, reviewing and editing the draft. All authors have read and approved the final manuscript.

## Data availability statement

The data that support the findings of this study are available from the corresponding author upon reasonable request.

## Abbreviations

SARS-CoV-2: Severe Acute Respiratory Syndrome Coronavirus 2
CVDs: Cardiovascular diseases
DRPIs: Device-related pressure injuries
MASD: moisture-associated skin damage
STs: skin tears
TNF: tumor necrosis factor
IL: interleukin
B: coefficient of regression *β* (beta)
SE: standard error
OR: odds ratio
95% CI: 95% confidence interval.
PPE: personal protective equipment

